# A machine learning model to aid detection of familial hypercholesterolaemia

**DOI:** 10.1101/2022.06.17.22276540

**Authors:** Jasmine Gratton, Marta Futema, Steve E. Humphries, Aroon D. Hingorani, Chris Finan, A. Floriaan Schmidt

**Author notes:** **Corresponding author:** Jasmine Gratton, MRes, Institute of Cardiovascular Science, UCL, 222 Euston Road, NW1 2DA, London, United Kingdom. Contributed equally (Chris Finan, A. Floriaan Schmidt).

## Abstract

2.

**Background and Aims:** People with monogenic familial hypercholesterolaemia (FH) are at an increased risk of premature coronary heart disease and death. Currently there is no population screening strategy for FH, and most carriers are identified late in life, delaying timely and cost-effective interventions. The aim was to derive an algorithm to improve detection of people with monogenic FH.

**Methods:** A penalised (LASSO) logistic regression model was used to identify predictors that most accurately identified people with a higher probability of FH in 139,779 unrelated participants of the UK Biobank, including 488 FH carriers. Candidate predictors included information on medical and family history, anthropometric measures, blood biomarkers, and an LDL-C polygenic score (PGS). Model derivation and evaluation was performed using a random split of 80% training and 20% testing data.

**Results:** A 14-variable algorithm for FH was derived, where the top five variables included triglyceride, LDL-C, and apolipoprotein A1 concentrations, self-reported statin use, and an LDL-C PGS. Model evaluation in the test data resulted in an area under the curve (AUC) of 0.77 (95% CI: 0.71; 0.83), and appropriate calibration (calibration-in-the-large: -0.07 (95% CI: -0.28; 0.13); calibration slope: 1.02 (95% CI: 0.85; 1.19)). Employing this model to prioritise people with suspected monogenic FH is anticipated to reduce the number of people requiring sequencing by 88% compared to a population-wide sequencing screen, and by 18% compared to prioritisation based on LDL-C and statin use.

**Conclusions:** The detection of individuals with monogenic FH can be improved with the inclusion of additional non-genetic variables and a PGS for LDL-C.

## 3. INTRODUCTION

Familial hypercholesterolaemia (FH) is an autosomal dominant disorder caused by variants in the *LDLR, APOB, PCSK9*, or *APOE* genes. It is characterised by elevated low-density lipoprotein (LDL-C) concentration and premature coronary heart disease (CHD).(1) FH-causing variants are found in about 1 in 250 individuals (95% CI: 1:345; 1:192),(2) however the condition remains highly underdiagnosed worldwide with only an estimated 1% to 10% of cases diagnosed.(3,4) Affected individuals are at increased risk of premature CHD, where early initiation of lipid-lowering treatment is paramount for risk management.(3) There is currently no systematic way of identifying new index FH cases in the general population, although cascade testing in families of affected individuals has been shown to be highly cost-effective in many countries.(5–8) Currently, patient diagnosis often happens after the development of CHD symptoms or by opportunistic measurement of lipid profile and at the discretion of clinicians. Diagnosis is made using tools such as the Dutch Lipid Clinical Network (DLCN) and the Simon Broome criteria, which have not been designed to be used as population screening tools.(1)

In 2016, Wald *et al*. suggested screening children aged 15 months of age by measurement of total or LDL-C to systematically identify index monogenic FH cases in the general population as a prelude to testing parents and other family members.(9) Futema *et al*. showed that measurement of LDL-C alone at age 9 may be insufficiently accurate in reliably distinguishing FH-variant carriers from those with an elevated cholesterol as a consequence diet and lifestyle factors, or carriage of a high burden of common cholesterol-raising alleles, and suggested adding a confirmatory targeted-sequencing step to reduce the number of false positive cases detected.(10)

The increased availability of routine health checks in adults either through work-place schemes or local healthcare providers offers an opportunity to systematically identify adult carriers of FH-causing variants.(11) Positioning adult FH screening within routine health checks, which typically record a substantial number of other clinical measurements, offers the opportunity to consider additional predictors for FH. This may be important because, while the effect of FH on CHD risk is mediated through elevated circulating LDL-C concentration, it is well-known that LDL-C concentration associates with other variables such as blood and liver biomarkers, diet, and also with common, genetic variants.(12) Combining multiple environmental factors and a polygenic score for LDL-C raising genetic variants may improve the detection of people with monogenic FH for prioritisation for confirmatory genetic testing.(13,14) This is because individuals with monogenic FH are likely to have a measured LDL-C concentration that is higher than can be accounted for by these other variables.

In the current manuscript we utilise the UK Biobank data to evaluate the detection rate and testing burden of three prioritisation strategies to identify people with suspected FH-causing variants for confirmatory genetic testing: 1) no prioritisation (i.e., referring all participants for sequencing), 2) a plasma LDL-C-based prioritisation model adjusting for statin treatment, 3) a multivariable machine learning prioritisation model.

## 4. METHODS

### Available genomics data and FH ascertainment

We identified 472,147 UK Biobank participants of White British ancestry (data-field 21000) as part of the approved project ID 40721. After performing genomic quality control steps (see Supplementary Material page 1), 341,515 individuals remained, including 140,439 with whole-exome sequencing (WES) data necessary to identify those who carry an FH-causing variant. Causal FH variants were searched for in the WES data encompassing the *LDLR, APOB, PCSK9* and *APOE* genes (Online Methods section of the Supplementary Material and Supplementary Table 1). A total of 488 pathogenic and likely pathogenic FH variants were identified (Supplementary Table 2). Additionally, 660 participants were found to carry FH variants of uncertain significance (VUS) (Supplementary Table 3). These were excluded from the analysis because more evidence is required to interpret the effect of those VUS.

### LDL-C PGS generation

We next generated a PGS for LDL-C concentration using an independent data subset of 173,672 White British participants without lipid-lowering medication or WES data (Supplementary Figure 1). An initial list of 10,137 genetic variants with a p-value threshold of <5×10^−4^ was obtained from the Global Lipids Genetics Consortium (GLGC) genome-wide association study (GWAS) summary statistics for LDL-C.(15) To reduce the number of potentially redundant variants and optimise LDL-C prediction, we next applied a least absolute shrinkage and selection operator (LASSO) regression algorithm using the biglasso package in R.(16) The degree of penalisation was determined through 15-fold cross-validation, maximising the explained variance (R-squared), which resulted in a 1,466 genetic variant LDL-C PGS.

### Deriving a machine learning algorithm to prioritise participants with FH

We extracted data on a total of 24 candidate FH predictors, specifically: LDL-C, high-density lipoprotein cholesterol (HDL-C), total cholesterol, triglycerides, lipoprotein A (Lp(a)), apolipoprotein A1 (Apo-A1), apolipoprotein B (Apo-B), C-reactive protein (CRP), aspartate aminotransferase (AST), alanine aminotransferase (ALT), alkaline phosphatase (ALP), sex, body mass index (BMI), age, self-reported statin use, alcohol use, systolic blood pressure (SBP), diastolic blood pressure (DBP), Townsend deprivation index, smoking status, family history of CHD, type 2 diabetes diagnosis, hypertension, and LDL-C PGS. This was expanded by including 10 product terms between: age and LDL-C, age and LDL-C PGS, LDL-C PGS and LDL-C, age^2^, LDL-C^2^, statin use and LDL-C, family history of CHD and sex, family history of CHD and statin use, family history of CHD and alcohol use, family history of CHD and hypertension. The limited missing data (Supplementary Table 4) were singly imputed using the R package MICE.(17)

Model derivation was performed using the WES data, applying a 80% training data split of 111,824 subjects, retaining 20% testing data (containing 93 carriers out of 27,955 subjects) to unbiasedly evaluate model performance (Supplementary Figure 1). To prevent potential model instability, highly correlated variables (i.e. multicollinear) were removed. These included Apo-B and total cholesterol (Supplementary Figure 2). Variables were standardised to mean zero and standard deviation (SD) one. Finally, we applied a binomial regression model with LASSO penalisation to derive a discrimination-optimised FH prediction model. Specifically, optimal penalisation was determined through 15-fold cross-validation maximising the c-statistic (i.e., the area under the receiver operating characteristic (AUC-ROC) curve).(16)

Model performance was evaluated using the 20% testing data based on its discriminative ability (c-statistic), appropriate calibration of predicted and observed probability of having an FH variant (using calibration plots, calibration-in-the-large, and calibration slope), and classification metrics (sensitivity, specificity (or its compliment the false positive rate), positive predictive value, and the negative predicted value).

### Evaluating the burden of genomic sequencing for FH

While genetic sequencing is the gold standard for FH diagnosis, it may often be prohibitively expensive to offer it to an entire population as a screening strategy. We therefore explored whether prioritising people with suspected FH can reduce the screening burden with an acceptable number of false-negative results. We evaluated the following prioritisation strategies: 1) no prioritisation (i.e. referring all participants for sequencing), 2) prioritisation based on LDL-C concentration (adjusting for statin use), 3) a multivariable model built from genetic, clinical biomarkers and environmental predictors.

These prioritisation strategies were evaluated on the number of subjects that would need to be sequenced, the proportion of FH carriers who would be missed, and the ratio of FH carriers correctly prioritised by the number of non-carriers unnecessarily offered sequencing. Additionally, prioritisation based on LDL-C concentrations (adjusted for statin use) was compared to prioritisation using the multivariable model with the help of a net reclassification index (NRI) analysis.

## 5. RESULTS

### Participant characteristics of our study cohort

Using the UK Biobank WES data, we identified 488 pathogenic or likely pathogenic FH variant carriers (list of variants shown in Supplementary Table 2) and 139,291 non-carriers; 0.35% (95% confidence interval (CI): 0.32; 0.38). FH variant carriers had a significantly higher frequency of a family history of coronary heart disease (CHD) (62.7% versus 48.1% in controls), higher prevalence (8.2% versus 2.8% in controls) and incidence (6.6% versus 3.9% in controls) of CHD (Supplementary Material and Table 1).

**Table 1.**
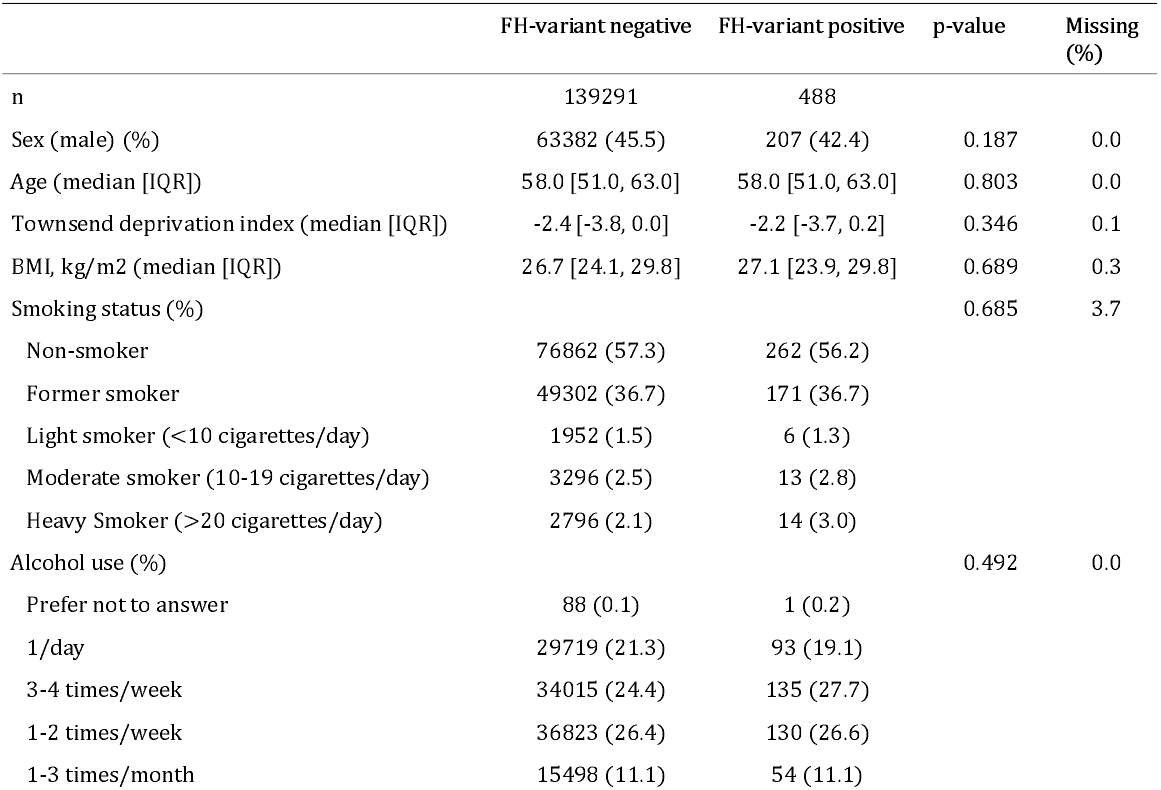
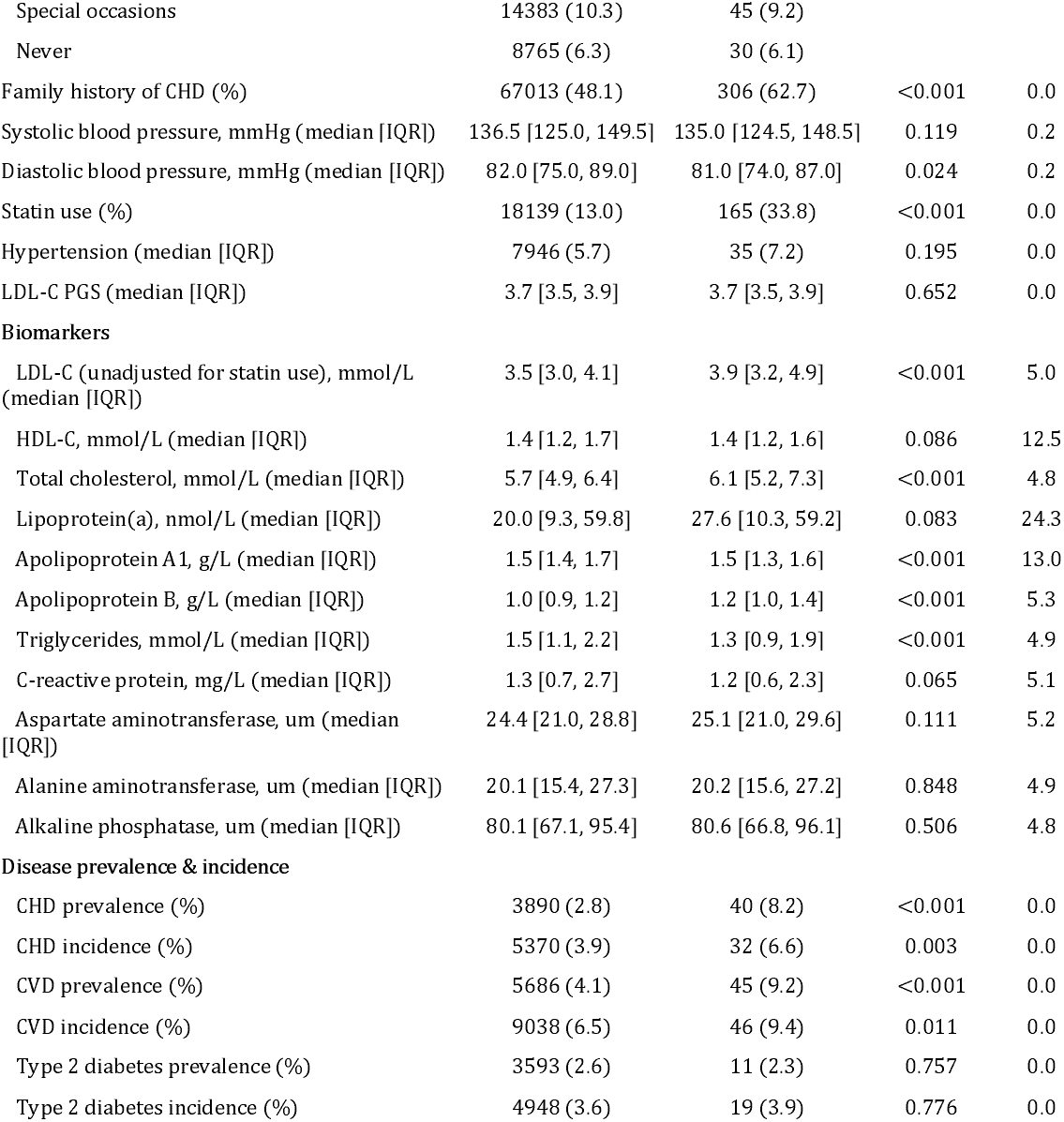
UK Biobank participant characteristics stratified by carrying a familial hypercholesterolaemia (FH)-causing variant. The p-values shown in the table are from the Kruskal-Wallis Rank Sum test for continuous variables, and from the Man-Whitney U test for binary variables. IQR = interquartile range; BMI = body mass index; CHD = coronary heart disease; LDL-C = low-density lipoprotein cholesterol; HDL-C = high-density lipoprotein cholesterol; CVD = cardiovascular disease; PGS = polygenic score.

|  | FH-variant negative | FH-variant positive | p-value | Missing (%) |
| --- | --- | --- | --- | --- |
| n | 139291 | 488 |  |  |
| Sex (male) (%) | 63382 (45.5) | 207 (42.4) | 0.187 | 0.0 |
| Age (median [IQR]) | 58.0 [51.0, 63.0] | 58.0 [51.0, 63.0] | 0.803 | 0.0 |
| Townsend deprivation index (median [IQR]) | -2.4 [-3.8, 0.0] | -2.2 [-3.7, 0.2] | 0.346 | 0.1 |
| BMI, kg/m<sup>2</sup> (median [IQR]) | 26.7 [24.1, 29.8] | 27.1 [23.9, 29.8] | 0.689 | 0.3 |
| Smoking status (%) |  |  | 0.685 | 3.7 |
| Non-smoker | 76862 (57.3) | 262 (56.2) |  |  |
| Former smoker | 49302 (36.7) | 171 (36.7) |  |  |
| Light smoker (<10 cigarettes/day) | 1952 (1.5) | 6 (1.3) |  |  |
| Moderate smoker (10-19 cigarettes/day) | 3296 (2.5) | 13 (2.8) |  |  |
| Heavy Smoker (>20 cigarettes/day) | 2796 (2.1) | 14 (3.0) |  |  |
| Alcohol use (%) |  |  | 0.492 | 0.0 |
| Prefer not to answer | 88 (0.1) | 1 (0.2) |  |  |
| 1/day | 29719 (21.3) | 93 (19.1) |  |  |
| 3-4 times/week | 34015 (24.4) | 135 (27.7) |  |  |
| 1-2 times/week | 36823 (26.4) | 130 (26.6) |  |  |
| 1-3 times/month | 15498 (11.1) | 54 (11.1) |  |  |
| Special occasions | 14383 (10.3) | 45 (9.2) |  |  |
| Never | 8765 (6.3) | 30 (6.1) |  |  |
| Family history of CHD (%) | 67013 (48.1) | 306 (62.7) | <0.001 | 0.0 |
| Systolic blood pressure, mmHg (median [IQR]) | 136.5 [125.0, 149.5] | 135.0 [124.5, 148.5] | 0.119 | 0.2 |
| Diastolic blood pressure, mmHg (median [IQR]) | 82.0 [75.0, 89.0] | 81.0 [74.0, 87.0] | 0.024 | 0.2 |
| Statin use (%) | 18139 (13.0) | 165 (33.8) | <0.001 | 0.0 |
| Hypertension (median [IQR]) | 7946 (5.7) | 35 (7.2) | 0.195 | 0.0 |
| LDL-C PGS (median [IQR]) | 3.7 [3.5, 3.9] | 3.7 [3.5, 3.9] | 0.652 | 0.0 |
| **Biomarkers** |  |  |  |  |
| LDL-C (unadjusted for statin use), mmol/L (median [IQR]) | 3.5 [3.0, 4.1] | 3.9 [3.2, 4.9] | <0.001 | 5.0 |
| HDL-C, mmol/L (median [IQR]) | 1.4 [1.2, 1.7] | 1.4 [1.2, 1.6] | 0.086 | 12.5 |
| Total cholesterol, mmol/L (median [IQR]) | 5.7 [4.9, 6.4] | 6.1 [5.2, 7.3] | <0.001 | 4.8 |
| Lipoprotein(a), nmol/L (median [IQR]) | 20.0 [9.3, 59.8] | 27.6 [10.3, 59.2] | 0.083 | 24.3 |
| Apolipoprotein A1, g/L (median [IQR]) | 1.5 [1.4, 1.7] | 1.5 [1.3, 1.6] | <0.001 | 13.0 |
| Apolipoprotein B, g/L (median [IQR]) | 1.0 [0.9, 1.2] | 1.2 [1.0, 1.4] | <0.001 | 5.3 |
| Triglycerides, mmol/L (median [IQR]) | 1.5 [1.1, 2.2] | 1.3 [0.9, 1.9] | <0.001 | 4.9 |
| C-reactive protein, mg/L (median [IQR]) | 1.3 [0.7, 2.7] | 1.2 [0.6, 2.3] | 0.065 | 5.1 |
| Aspartate aminotransferase, um (median [IQR]) | 24.4 [21.0, 28.8] | 25.1 [21.0, 29.6] | 0.111 | 5.2 |
| Alanine aminotransferase, um (median [IQR]) | 20.1 [15.4, 27.3] | 20.2 [15.6, 27.2] | 0.848 | 4.9 |
| Alkaline phosphatase, um (median [IQR]) | 80.1 [67.1, 95.4] | 80.6 [66.8, 96.1] | 0.506 | 4.8 |
| **Disease prevalence & incidence** |  |  |  |  |
| CHD prevalence (%) | 3890 (2.8) | 40 (8.2) | <0.001 | 0.0 |
| CHD incidence (%) | 5370 (3.9) | 32 (6.6) | 0.003 | 0.0 |
| CVD prevalence (%) | 5686 (4.1) | 45 (9.2) | <0.001 | 0.0 |
| CVD incidence (%) | 9038 (6.5) | 46 (9.4) | 0.011 | 0.0 |
| Type 2 diabetes prevalence (%) | 3593 (2.6) | 11 (2.3) | 0.757 | 0.0 |
| Type 2 diabetes incidence (%) | 4948 (3.6) | 19 (3.9) | 0.776 | 0.0 |

### Multivariable machine learning model to prioritise FH variant carriers

14 out of the 32 variables were retained by the LASSO regression model for the prediction of FH (Figure 1.a, Supplementary Figure 3, Supplementary Table 5), including triglyceride, Apo-A1, ALT and CRP concentrations, statin use, LDL-C PGS, family history of CHD, DBP, BMI, and prevalent T2D. Additionally, the following product terms were selected: LDL-C^2^, statin use and LDL-C, age and LDL-C PGS. Retention of these product terms indicated the presence of non-linear associations with FH, for example the LDL-C association with the presence of a monogenic FH variant was found to be quadratic (Supplementary Figure 4).

**Figure 1.**
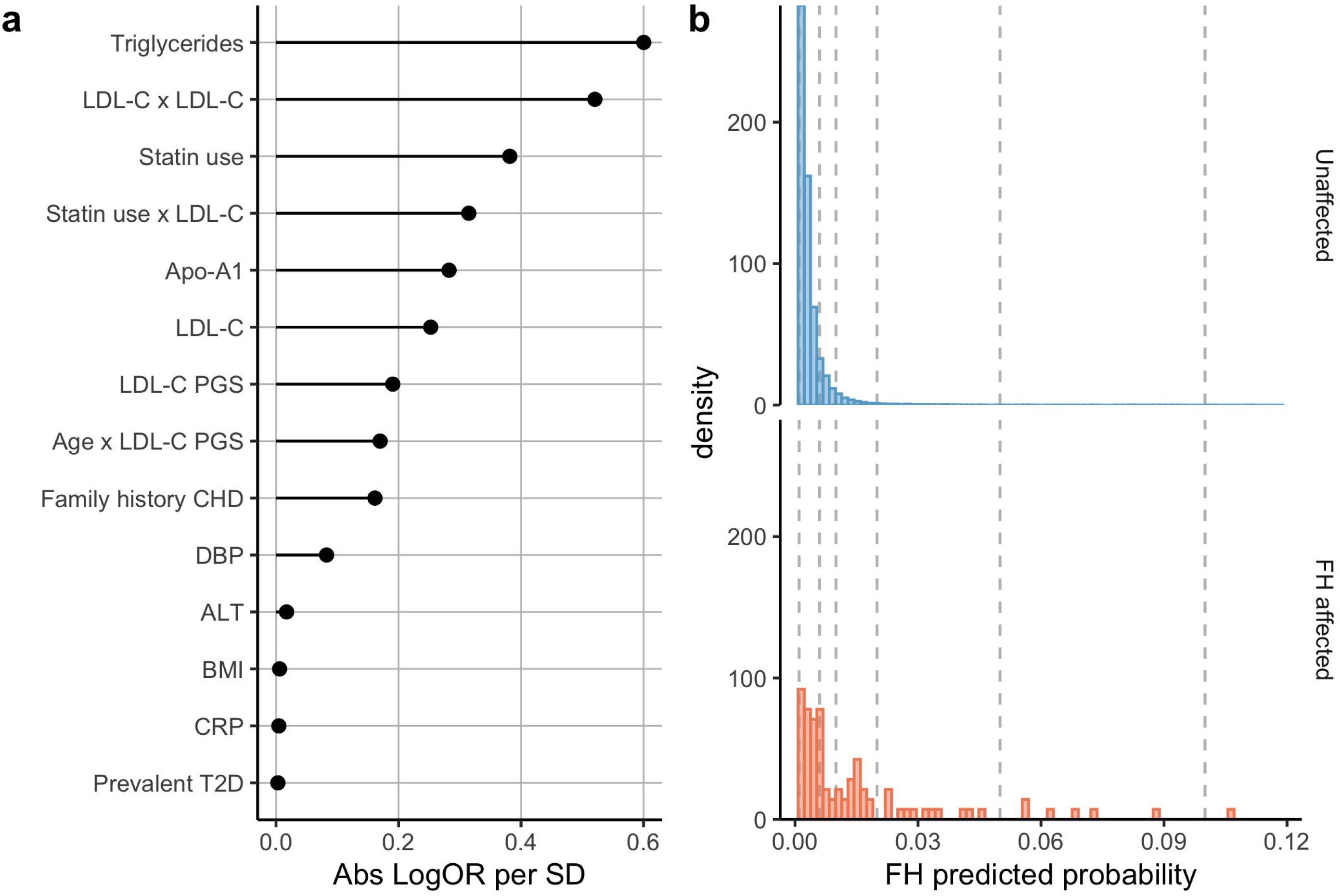
Feature importance of the variables retained by LASSO regression predicting monogenic FH, and the density predicted probability distributions from this model for unaffected and affected FH individuals in White British participants of the UK Biobank. **a)** The 14 predictors retained by LASSO regression ordered by absolute log odds ratio (OR) per standard deviation (SD). The “x” sign is used to indicate an interaction term. Abs = absolute; LDL-C = low-density lipoprotein cholesterol; Apo-A1 = apolipoprotein A1; PGS = polygenic score; CHD = coronary heart disease; DBP = diastolic blood pressure; ALT = alanine aminotransferase; BMI = body mass index; CRP = C-reactive protein; T2D = type 2 diabetes. **b)** The density predicted probability distributions for affected (in orange) and unaffected (in blue) FH participants in our test cohort as predicted by the multivariable model. 14 unaffected individuals had a monogenic FH predicted probability above 0.12 and are not shown on the plot for legibility purposes. The vertical dotted lines represent the various FH predicted probability thresholds evaluated in Table 2.

The median predicted probability of having monogenic FH by the multivariable model was ∼3 fold higher in FH carriers (0.64%, interquartile range (IQR): 0.31; 1.62) compared to non-carriers (0.23%, IQR: 0.14; 0.38), with partial overlap between FH carriers and non-carriers (Figure 1.b). The test data AUC for this model was 0.77 (95% CI: 0.71; 0.83), with a training data AUC of 0.78 (95% CI: 0.75; 0.81). Calibration statistics (calibration-in-the-large: -0.073 (95% CI: -0.28; 0.13) and calibration slope: 1.02 (95% CI: 0.85; 1.19)) indicated the predicted probability agreed well with the observed probability (Figure 2.a).

**Table 2.** The classification accuracy of an algorithm for predicting monogenic familial hypercholesterolaemia (FH) using the multivariable model and LDL-C concentration accounting for statin use. There are 93 FH-causing variant positive participants in the test data comprising of a total of 27,955 participants.

| **Predicted probability cut-off** | **% sensitivity (95%CI)** | **% false positive rate (95%CI)** | **% positive predictive value (95%CI)** | **% negative predictive value (95%CI)** | **FH-causing variants below threshold** | **FH-causing variants above threshold** | **Controls above threshold** |
| --- | --- | --- | --- | --- | --- | --- | --- |
| **Multivariable model** |  |  |  |  |  |  |  |
| **0.1** | 1.1 (0.2;5.8) | 0.1 (0.0;0.1) | 5.6 (1.0;25.8) | 99.7 (99.6;99.7) | 92 | 1 | 17 |
| **0.05** | 7.5 (3.7;14.7) | 0.2 (0.1;0.2) | 13.0 (6.4;24.4) | 99.7 (99.6;99.8) | 86 | 7 | 47 |
| **0.02** | 20.4 (13.5;29.7) | 1.1 (0.9;1.2) | 6.0 (3.9;9.2) | 99.7 (99.7;99.8) | 74 | 19 | 296 |
| **0.01** | 41.9 (32.4;52.1) | 4.5 (4.2;4.7) | 3.0 (2.2;4.1) | 99.8 (99.7;99.8) | 54 | 39 | 1244 |
| **0.006** | 54.8 (44.7;64.6) | 11.9 (11.5;12.3) | 1.5 (1.2;2.0) | 99.8 (99.8;99.9) | 42 | 51 | 3311 |
| **0.001** | 94.6 (88.0;97.7) | 87.0 (86.6;87.4) | 0.4 (0.3;0.4) | 99.9 (99.7;99.9) | 5 | 88 | 24240 |
| **Model: LDL-C concentration + statin use** |  |  |  |  |  |  |  |
| **0.1** | 0.0 (0.0;4.0) | 0.0 (0.0;0.1) | 0.0 (0.0;35.4) | 99.7 (99.6;99.7) | 93 | 0 | 7 |
| **0.05** | 1.1 (0.2;5.8) | 0.1 (0.1;0.2) | 3.2 (0.6;16.2) | 99.7 (99.6;99.7) | 92 | 1 | 30 |
| **0.02** | 12.9 (7.5;21.2) | 1.1 (1.0;1.2) | 3.8 (2.2;6.5) | 99.7 (99.6;99.8) | 81 | 12 | 304 |
| **0.01** | 38.7 (29.4;48.9) | 5.6 (5.4;5.9) | 2.2 (1.6;3.1) | 99.8 (99.7;99.8) | 57 | 36 | 1574 |
| **0.006** | 52.7 (42.6;62.5) | 14.6 (14.2;15.0) | 1.2 (0.9;1.6) | 99.8 (99.8;99.9) | 44 | 49 | 4067 |
| **0.001** | 90.3 (82.6;94.8) | 84.0 (83.5;84.4) | 0.4 (0.3;0.4) | 99.8 (99.6;99.9) | 9 | 84 | 23393 |

**Figure 2.**
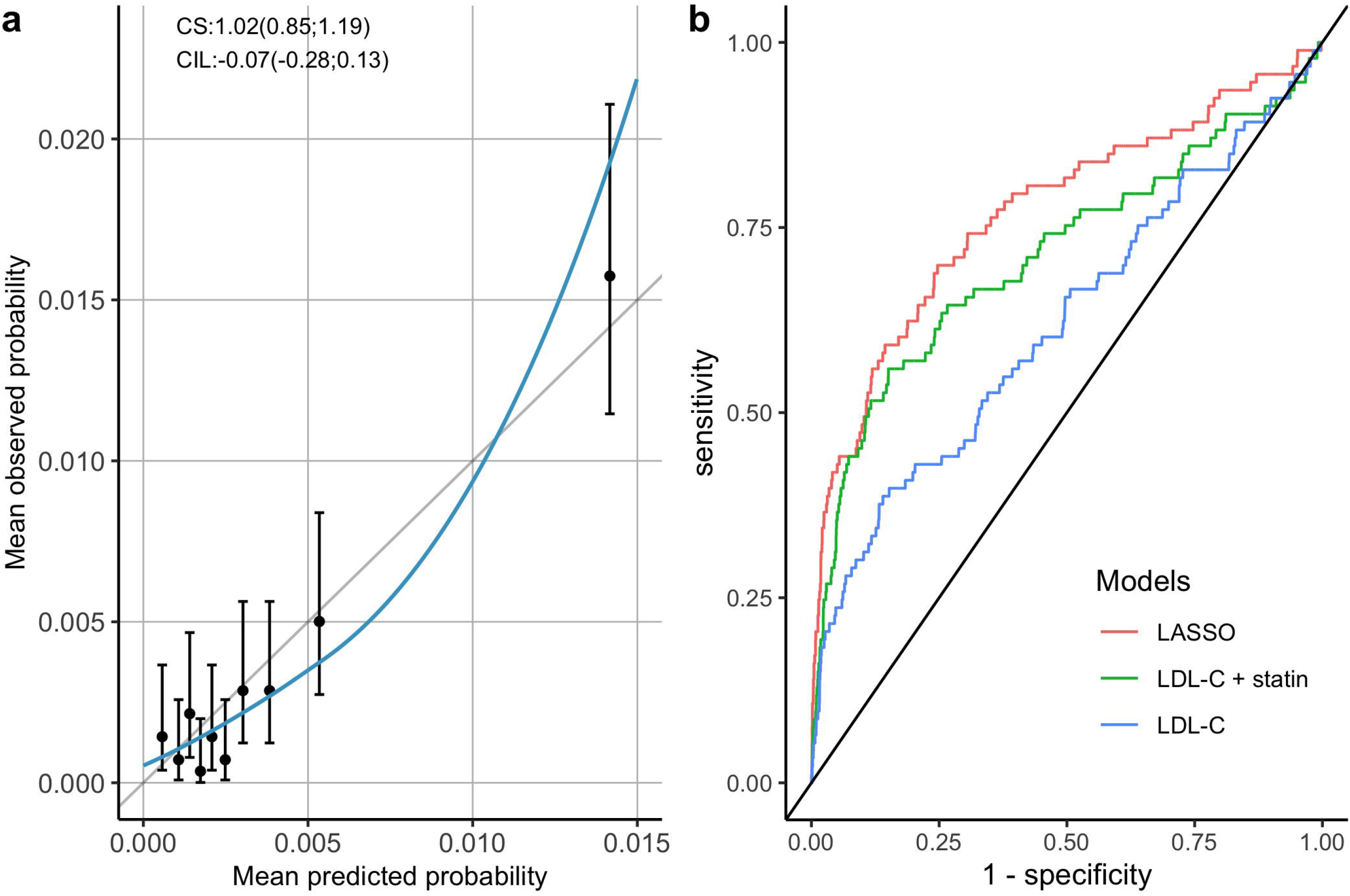
Discrimination and calibration of a multivariable algorithm predicting FH carriership using independent testing data. **a)** The calibration plot for the multivariable model where the mean predicted and mean observed probability for each decile of the test data are depicted by the datapoints with their 95% confidence intervals (CI). Perfect calibration is indicated by the vertical black line. The calibration-in-the-large (CIL) and the calibration slope (CS) values are indicated on the plot with their 95% CI in brackets. The loess line was fitted with FH-causing variant status as the outcome and mean predicted probability as the predictor. **b)** The receiver operating characteristic (ROC) curves for the multivariable model (in red), LDL-C concentration and statin model (in green), and LDL-C concentration only model (in blue). The area under the curve (AUC) for each of these models are equal to 0.77 (95% CI: 0.71; 0.83), 0.71 (95% CI: 0.65; 0.77) and 0.62 (95% CI: 0.56; 0.68) respectively.

The multivariable machine learning model outperformed a model which only consider LDL-C (AUC: 0.62, 95% CI: 0.56; 0.68), as well as a model which additionally included a statin indicator (AUC: 0.71, 95% CI: 0.65; 0.77), both evaluated in the test data. (Figure 2.b).

### Model FH classification

Next, we evaluated the FH classification performance of the multivariable model using six cut-off values of having an FH variant (from 0.001 to 0.10) in the test dataset. The sensitivity increased from 1.1% (95% CI: 0.2; 5.8) for a predicted probability of 0.10, to 94.6% (95% CI: 88.0; 97.7) for a predicted probability of 0.001; with the false positive rate similarly increasing from 0.1% (95% CI: 0.0; 0.1) to 87.0% (95% CI: 86.6; 87.4) (Table 2). We further compared the performance of these thresholds to a simpler model of LDL-C concentration adjusted for statin, which underperformed (Table 2).

The net reclassification index (NRI) comparing the LDL-C and statin use model to the multivariable model, indicated that the improved performance of the latter was due to it assigning a higher predicted probability to FH variant carriers. At a predicted probability threshold of 0.006, the probability for FH carriers being reclassified as having an FH variant was equal to 0.097 (95% CI: 0.038; 0.159), as opposed to the probability of 0.075 (95% CI: 0.026; 0.130) of being down-classified as not having an FH variant (Table 3).

**Table 3.**
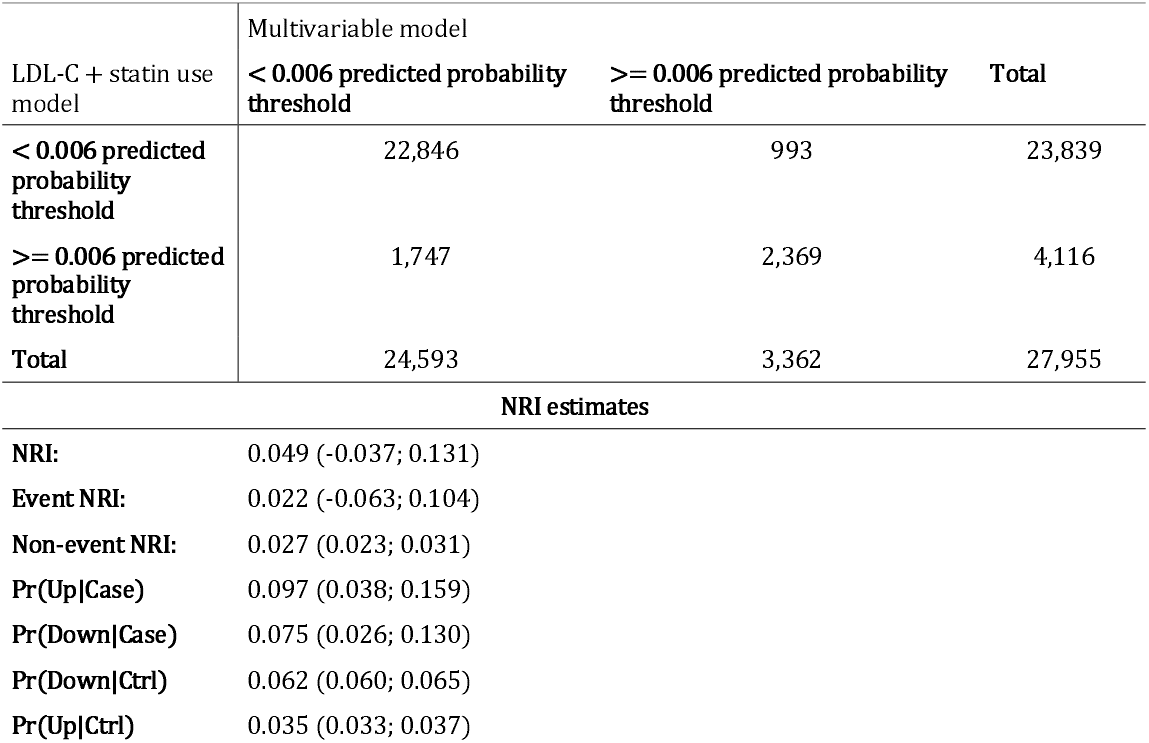
Net reclassification index (NRI) table and estimates for a predicted probability threshold of 0.006 comparing the multivariable model to a simpler model of LDL-C concentration and statin use. The predicted probability threshold of 0.006 was chosen to illustrate the NRI analysis between the multivariable model and the LDL-C with statin use model. This threshold choice was chosen as it had a false positive rate close to 10% (11.9%). The test dataset of 27,955 participants was used, which included 93 FH variant carriers. NRI estimates were obtained via percentile bootstrap method. Pr = probability; Up = reclassified to the higher category; Down = reclassified to the lower category; Case = FH-causing variant positive; Ctrl = control (negative for an FH-causing variant).

### Prioritising individuals for FH genomic testing in a two-stage population screening strategy

Finally, we evaluated the performance of a two-stage population screen for identifying new index FH cases, were the second stage consisted of targeted sequencing of FH variants (Supplementary Figure 5). The multivariable and LDL-C with statin use models were compared using a common threshold of 0.006, where on average, seven additional FH carriers would be detected for 100,000 individuals screened when using the multivariable model compared to the LDL-C and statin use model. Per 100,000 individuals screened, the multivariable model would refer 12,033 individuals (12%) for genetic sequencing, compared to 14,730 (15%) with the LDL-C and statin use model, resulting in a 18% reduction in genetic testing.

Furthermore, if we assume that FH has a population prevalence of 1 in 286 (equal to our cohort’s prevalence) and that one FH case has on average 1.5 first-degree relatives ((2 children + 1 sibling) / 2) who are also affected by FH (discovered through cascade testing),(18) then overall one FH case would be identified for every ∼219 people screened when using the multivariable model, compared to one FH case for every ∼228 individuals screened with the LDL-C and statin use model.

## 6. DISCUSSION

In the current manuscript we derived a multivariable machine learning model to identify people with suspected FH for confirmatory DNA sequencing in the context of population screening. Using LASSO regression, we derived a 14-feature model consisting of LDL-C, Apo-A1, triglyceride, ALT, and CRP concentrations, self-reported statin use, family history of CHD, DBP, BMI, type 2 diabetes diagnosis, three product terms, and an LDL-C PGS. The multivariable algorithm was able to discriminate between FH and non-FH carriers with an AUC of 0.77 (95% CI: 0.71; 0.83), with good calibration, outperforming a simpler model consisting of LDL-C and an indicator for statin prescription.

Independent of the classification threshold applied, the multivariable algorithm was able to substantially decrease the number of subjects referred to genetic sequencing (e.g. from 100,000 individuals without any prioritisation, to 14,730 with prioritisation using the LDL-C and statin use model, and to 12,033 with prioritisation using the multivariable model for a predicted probability threshold of carrying a variant for monogenic FH of 0.006; equivalent to approximately a 18% decrease in individuals needed to be sequenced between the last two models). These differences become especially significant if extrapolating the values to a population-wide scale comprising of millions of participants screened. Our results provide support for opportunistic screening and seeding of cascade testing for FH, which could be integrated within existing health checks offered to employers or local healthcare providers.(11)

Previously, Banda *et al*. used a machine learning method to detect monogenic FH cases from electronic health records (EHR).(19) While their model showed an impressive AUC of 0.94, one of their most important features was referral to a cardiology clinic, which is in very close proximity to confirmatory FH testing, limiting the model’s utility as a prospective tool for FH diagnosis. Besseling *et al*. developed a multivariable model to identify FH carriers validated in study participants consisting of FH cases and their relatives, again limiting applicability to the general population.(20) Our model instead considers FH prioritisation in a non-GP-referred population and is more generalisable as a systematic population screening tool.

Our multivariable model included three terms for LDL-C (LDL-C itself, LDL-C squared, and an interaction with statin prescription), which combined makes it the most important predictor. Additionally, our model also identified novel predictors for FH such as triglyceride and Apo-A1 concentrations, with triglycerides having the largest absolute OR per SD (0.60). High triglyceride levels are often linked to poor diet and a sedentary lifestyle.(21– 24) Here we find that FH carriers had lower triglyceride concentrations than non-carriers (Table 1), which resulted in a protective association, indicating that triglyceride concentrations can be useful in discriminating between individuals who have hypercholesterolaemia due to lifestyle factors as opposed to an FH-causing variant. We also found that higher Apo-A1 concentrations, a protein found on HDL particles, was associated with a decreased probability of FH. Finally, we note that our multivariable FH model retained a squared term for LDL-C, suggesting that LDL-C is not linearly related with carrying an FH variant, but rather has a quadratic relationship (Supplementary Figure 4).

The variables included in our multivariable algorithm should not be interpreted as causal risk factors for monogenic FH; they simply help to distinguish non-monogenetic sources of variation in LDL-C concentrations from monogenic causes (as was discussed in more detail previously with triglyceride concentrations). This also provides the rational for including an LDL-C PGS in the model: a large discrepancy between predicted LDL-C concentrations (by the LDL-C PGS) and observed LDL-C concentrations might be indicative of FH carriership,(13,14) demonstrated here by a negative coefficient for LDL-C PGS in the model (Supplementary Table 5). We note that a previous LDL-C PGS by Wu *et al*. had a substantially larger R-squared (0.21 (95% CI: 0.20-0.22)) than reported here (0.14 (95% CI: 0.13-0.15)).(25) Unlike Wu *et al*. who identified genetic variants from an internal UK Biobank LDL-C GWAS overlapping with the PGS training data; we identified variants based on an independent dataset from GLGC,(15) guarding against overfitting through ‘data-leakage’ between the training and testing datasets and providing a more robust estimate of explained variance.

A study limitation to consider is the exclusion of individuals with VUS from our study cohort. There is conflicting evidence as to the causal effects of these VUS in FH. We anticipate that some are likely to be FH-causing while others are not, but more research is needed. As more VUS are classified as either FH-causing or not, the model can be readily updated to reflect our growing understanding of FH. Additionally, it is impossible to know whether some study participants have been genetically tested for carrying an FH variant, and whether they might have modified their behaviour (e.g. diet) following their diagnosis. This could potentially impact the accuracy of the multivariable model developed here; however, considering that only approximately 7% of FH cases have been diagnosed in the UK,(26) this low number of diagnoses is unlikely to have a significant effect on the model and results presented here. Currently, PGS information is not routinely used or collected in clinical practice, which is why we also derived a penalised multivariable model without an LDL-C PGS, which did not meaningfully decrease performance (Supplementary Table 6). Previous studies have suggested that PGS could be used to identify individuals with a rare variant for certain diseases, such as FH.(13,14) Our study confirms the utility of the PGS for FH prioritisation; however given its correlation with environmental variables (e.g. lipid levels), this genetic information can be readily replaced with information from non-genetic data.

We have tested our multivariable model in a dataset which was independent from the training data, with no significant difference between training and testing AUC (difference of 0.01), suggesting limited model overfitting to the current sample. Nevertheless, considering the health discrepancies observed between the UK Biobank and the general UK population,(27) we suggest that this model is locally validated and updated before applying it to distinct settings. Model validation should especially be conducted when considering populations of non-European ancestry. Irrespective of the important considerations regarding model transferability, prior to integrating the model in clinical care, an informed decision should be made on the optimal predicted probability threshold for monogenic FH classification. We wish to highlight that our choice of 0.006 as a threshold is purely pragmatic, and a more optimised threshold could further increase benefit. Given that monogenic FH is relatively rare in the general population, we would expect the optimum probability threshold to be low, similar to the one employed here. While Youden’s J statistic can be used to identify the optimal threshold balancing sensitivity and specificity, this implies equal costs between false-positive and false-negative predictions which is unlikely to be true. The choice of threshold should be supported by (local) health-technology assessments incorporating direct and indirect costs.

In conclusion, we derived a multivariable classification model for detecting monogenic FH variant carriers that outperformed a model based on LDL-C concentration (adjusted for statin use) for FH screening, and that offers an opportunity to prioritise suspected FH carriers for genetic sequencing.

## Data Availability

All data produced in the present study are available upon reasonable request to the authors.

## 8. ACKNOWLEDGEMENTS

This research has been conducted using data from UK Biobank (application number 40721), a major biomedical database.(28) The authors are grateful to UK Biobank participants. UK Biobank was established by the Wellcome Trust medical charity, Medical Research Council, Department of Health, Scottish Government, and the Northwest Regional Development Agency. It has also had funding from the Welsh Assembly Government and the British Heart Foundation.

