## Supplementary Material for "A machine learning model to aid detection of familial hypercholesterolaemia"

**ONLINE METHODS**

**Quality control of the genotyping data**

Genotyped individuals of European ancestry (data-field 22006) of the UK Biobank (application number 40721) underwent stringent quality control (QC) prior to analysis. Outliers for heterozygosity and missingness (data-field 22027) and putative chromosomal aneuploidy (data-field 22019) were excluded from the dataset. Individuals with sex mismatches between their self-reported sex and their genetic sex, and participants with missing genetic sex information were also excluded. Variant QC of the imputed genotype data was performed with bgenix,^1^ and all genetic variants with an imputation information (INFO) score <0.3 were removed. For the QC of the remaining genetic variants, Bristol’s MRC-IEU protocol was followed: genetic variants were excluded if they had a minor allele frequency (MAF) <0.1%, MAF <0.5% and INFO <0.9, MAF <1% and INFO <0.8, MAF <3% and INFO <0.6.^2^ Related individuals (up to and including third degree relatives) were also excluded from the analysis.

**FH case detection in the UK Biobank**

Multiallelic sites were converted to biallelic sites using BCFtools version 1.11.^3^, and genetic variants were annotated using Ensembl’s Variant Effect Predictor (VEP) release 103.1.^4^ The *LDLR* variants in the canonical transcript ENST00000558518 were filtered for a minor allele frequency (MAF) of 0.0006, which is equal to the frequency of the most common FH variant (p.Arg3527Gln in *APOB*) according to the gnomAD database version 2.1.1.^5^ Further variant filtering steps included a minimum read depth of 10 and genotype quality of 20. The SAMtools plugin split-vep was used to keep variants that had a predicted consequence of missense or worse, and the resulting variants with a SIFT annotation of “tolerated” or a PolyPhen annotation of “benign” were excluded.^3,6,7^ These filtering steps were followed by manual curation of the variants by two expert reviewers (M.F. and S.E.H.) who respected the Association for Clinical Genomics Science (ACGS) guidelines and the evidence accrued from the LOVD database for *LDLR*.^8,9^ For the *APOE* gene, the heterozygous p.Leu167del in-frame deletion was considered to be FH-causing, and the pathogenic variants in *PCSK9* and *APOB* were filtered based on a pre-defined list of curated variants with functional assay backing.^10,11^

**CHD and CVD definition codes**

CHD was defined with the International Classification of Diseases 10 (ICD-10) codes I21, I22, I23, I24, I25.0, I25.1, I25.2, I25.3, I25.5, 125.6, I25.8, 125.9, I48, I50, I11.0, I13.0, I13.2, I32.2, I61, I63 and the Office of Population Censuses and Surveys Classification of Interventions and Procedures 4 (OPCS-4) codes K40, K41, K42, K43, K44.1, K44.8, K44.9, K45.1, K45.2, K45.3, K45.4, K45.5, K45.8, K45.9, K46.1, K46.2, K46.3, K46.4, K46.8, K46.9, K47.1, K49, K50, K75, K52.1, K57.1, K57.5, K62.1, K62.2, K62.3, K62.4, K62.5, X50.1, X50.2. CVD was defined with the ICD-10 codes I21, I22, I23, I24, I25.0, I25.1, I25.2, I25.3, I25.5, 125.6, I25.8, 125.9 and the OPCS-4 codes K40, K41, K42, K43, K44.1, K44.8, K44.9, K45.1, K45.2, K45.3, K45.4, K45.5, K45.8, K45.9, K46.1, K46.2, K46.3, K46.4, K46.8, K46.9, K47.1, K49, K50, K75.

**Software**

The data analysis and figures were done in R version 4.0.2.^12^ The R package tableone version 0.12.0. was used to make Table 1.^13^ The receiver operating characteristic (ROC) curves were plotted with the R package pROC version 1.16.2.^14^ The NRI analysis was done using the R package nricens version 1.6.^15^

11.	Lázaro, C., Lerner-Ellis, J. & Spurdle, A. *Clinical DNA variant interpretation theory and practice*. (Academic Press, 2021).

12.	R: The R Project for Statistical Computing. Available at: https://www.r-project.org/. (Accessed: 11th February 2022)

13.	Panos, A. & Mavridis, D. TableOne: an online web application and R package for summarising and visualising data. *Evid. Based. Ment. Health* **23**, 127–130 (2020).

14.	Robin, X. *et al.* pROC: An open-source package for R and S+ to analyze and compare ROC curves. *BMC Bioinformatics* **12**, 1–8 (2011).

15.	Inoue, E. nricens: NRI for Risk Prediction Models with Time to Event and Binary Response Data. (2018).

**SUPPLEMENTARY RESULTS**

**LDL-C polygenic score (PGS)**

The p-value cut-off of 5x10^-4^ for the variants of GLGC’s LDL-C genome-wide association study (GWAS) summary statistics included 10,137 variants. After running the LASSO regression on these variants using the LASSO training dataset described in the methods section, 1,466 genetic variants were retained by the model. The LDL-C PGS r-squared was 0.14 (95% confidence intervals (CI): 0.13-0.15) in the independent test data.

**SUPPLEMENTARY TABLES AND FIGURES**

**Supplementary Table 1. Genetic coordinates of FH-causing genes.** Genetic coordinates are mapped to GRCh38.

| **Gene name** | **Chromosome number** | **Start coordinate** | **End coordinate** |
| --- | --- | --- | --- |
| *LDLR* | 19 | 11,089,262 | 11,133,820 |
| *APOB* | 2 | 21,001,429 | 21,044,073 |
| *APOE* | 19 | 44,905,791 | 44,909,393 |
| *PCSK9* | 1 | 55,039,347 | 55,064,852 |

**Supplementary Table 2. Autosomal dominant FH-causing mutation identified in our study cohort.** Genetic coordinates are mapped to GRCh38.

| **Gene** | **Chromosome** | **Position** | **Reference allele** | **Alternate allele** | **Nucleotide change** | **Protein** | **Number of carriers** | **UKB frequency (1/n)** |
| --- | --- | --- | --- | --- | --- | --- | --- | --- |
| *APOB* | 2 | 21006289 | G | A | c.10579C>T | p.Arg3527Trp | 2 | 70,220 |
|  |  | 21006288 | C | T | c.10580G>A | p.Arg3527Gln | 99 | 1,419 |
| *APOE* | 19 | 44908791 | GCTC | G | c.499_501del | p.Leu167del | 13 | 10,803 |
| *LDLR* | 19 | 11100236 | C | G | c.81C>G | p.Cys27Trp | 1 | 140,439 |
|  |  | 11100291 | T | G | c.136T>G | p.Cys46Gly | 1 | 140,439 |
|  |  | 11100294 | G | A | c.139G>A | p.Asp47Asn | 5 | 28,088 |
|  |  | 11102705 | C | T | c.232C>T | p.Arg78Cys | 13 | 10,803 |
|  |  | 11102714 | C | T | c.241C>T | p.Arg81Cys | 2 | 70,220 |
|  |  | 11102732 | T | G | c.259T>G | p.Trp87Gly | 6 | 23,407 |
|  |  | 11102741 | G | A | c.268G>A | p.Asp90Asn | 5 | 28,088 |
|  |  | 11102765 | G | A | c.292G>A | p.Gly98Ser | 10 | 14,044 |
|  |  | 11102774 | G | A | c.301G>A | p.Glu101Lys | 12 | 11,703 |
|  |  | 11102787 | G | A | c.313+1G>A | . | 5 | 28,088 |
|  |  | 11102787 | G | C | c.313+1G>C | . | 1 | 140,439 |
|  |  | 11102787 | G | GT | c.313+2dup | . | 2 | 70,220 |
|  |  | 11105249 | C | T | c.343C>T | p.Arg115Cys | 2 | 70,220 |
|  |  | 11105268 | G | T | c.362G>T | p.Cys121Phe | 2 | 70,220 |
|  |  | 11105324 | G | A | c.418G>A | p.Glu140Lys | 1 | 140,439 |
|  |  | 11105339 | GTGCTCACCTGTGGTCCCGCCAGC | G | c.435_457del | p.Leu146ProfsTer26 | 1 | 140,439 |
|  |  | 11105407 | C | A | c.501C>A | p.Cys167Ter | 2 | 70,220 |
|  |  | 11105408 | G | A | c.502G>A | p.Asp168Asn | 14 | 10,031 |
|  |  | 11105415 | AC | A | c.513del | p.Asp172ThrfsTer34 | 1 | 140,439 |
|  |  | 11105448 | C | G | c.542C>G | p.Pro181Arg | 2 | 70,220 |
|  |  | 11105549 | C | T | c.643C>T | p.Arg215Cys | 4 | 35,110 |
|  |  | 11105567 | G | A | c.661G>A | p.Asp221Asn | 2 | 70,220 |
|  |  | 11105568 | A | G | c.662A>G | p.Asp221Gly | 5 | 28,088 |
|  |  | 11105585 | GAC | G | c.680_681del | p.Asp227GlyfsTer12 | 4 | 35,110 |
|  |  | 11105585 | GAC | GAG | c.681delinsG | p.Asp227Glu | 2 | 70,220 |
|  |  | 11105588 | G | T | c.682G>T | p.Glu228Ter | 2 | 70,220 |
|  |  | 11105589 | AG | A | c.685del | p.Glu229LysfsTer36 | 1 | 140,439 |
|  |  | 11106579 | C | T | c.709C>T | p.Arg237Cys | 1 | 140,439 |
|  |  | 11106588 | G | A | c.718G>A | p.Glu240Lys | 20 | 7,022 |
|  |  | 11106592 | T | C | c.722T>C | p.Phe241Ser | 1 | 140,439 |
|  |  | 11106631 | A | C | c.761A>C | p.Gln254Pro | 1 | 140,439 |
|  |  | 11107432 | C | A | c.858C>A | p.Ser286Arg | 1 | 140,439 |
|  |  | 11107433 | G | A | c.859G>A | p.Gly287Ser | 4 | 35,110 |
|  |  | 11107436 | G | A | c.862G>A | p.Glu288Lys | 1 | 140,439 |
|  |  | 11107461 | G | A | c.887G>A | p.Cys296Tyr | 1 | 140,439 |
|  |  | 11107481 | C | T | c.907C>T | p.Arg303Trp | 2 | 70,220 |
|  |  | 11107486 | C | G | c.912C>G | p.Asp304Glu | 4 | 35,110 |
|  |  | 11107512 | G | A | c.938G>A | p.Cys313Tyr | 2 | 70,220 |
|  |  | 11110660 | G | A | c.949G>A | p.Glu317Lys | 35 | 4,013 |
|  |  | 11110678 | G | A | c.967G>A | p.Gly323Ser | 1 | 140,439 |
|  |  | 11110714 | G | A | c.1003G>A | p.Gly335Ser | 3 | 46,813 |
|  |  | 11110738 | G | A | c.1027G>A | p.Gly343Ser | 8 | 17,555 |
|  |  | 11110759 | C | T | c.1048C>T | p.Arg350Ter | 4 | 35,110 |
|  |  | 11110760 | G | C | c.1049G>C | p.Arg350Pro | 4 | 35,110 |
|  |  | 11111571 | G | A | c.1118G>A | p.Gly373Asp | 1 | 140,439 |
|  |  | 11111619 | C | T | c.1166C>T | p.Thr389Met | 8 | 17,555 |
|  |  | 11113286 | G | A | c.1195G>A | p.Ala399Thr | 1 | 140,439 |
|  |  | 11113287 | C | A | c.1196C>A | p.Ala399Asp | 1 | 140,439 |
|  |  | 11113292 | CTCTTC | CTCT | c.1205_1206del | p.Phe403HisfsTer37 | 1 | 140,439 |
|  |  | 11113307 | C | T | c.1216C>T | p.Arg406Trp | 5 | 28,088 |
|  |  | 11113308 | G | A | c.1217G>A | p.Arg406Gln | 4 | 35,110 |
|  |  | 11113313 | G | A | c.1222G>A | p.Glu408Lys | 1 | 140,439 |
|  |  | 11113322 | A | G | c.1231A>G | p.Lys411Glu | 1 | 140,439 |
|  |  | 11113329 | C | T | c.1238C>T | p.Thr413Met | 14 | 10,031 |
|  |  | 11113337 | C | T | c.1246C>T | p.Arg416Trp | 2 | 70,220 |
|  |  | 11113419 | G | C | c.1328G>C | p.Trp443Ser | 1 | 140,439 |
|  |  | 11113426 | C | G | c.1335C>G | p.Asp445Glu | 5 | 28,088 |
|  |  | 11113554 | CA | C | c.1379del | p.His460ProfsTer47 | 1 | 140,439 |
|  |  | 11113590 | G | T | c.1414G>T | p.Asp472Tyr | 6 | 23,407 |
|  |  | 11113608 | G | A | c.1432G>A | p.Gly478Arg | 2 | 70,220 |
|  |  | 11113612 | T | C | c.1436T>C | p.Leu479Pro | 2 | 70,220 |
|  |  | 11113620 | G | A | c.1444G>A | p.Asp482Asn | 29 | 4,843 |
|  |  | 11113650 | G | A | c.1474G>A | p.Asp492Asn | 1 | 140,439 |
|  |  | 11113678 | C | T | c.1502C>T | p.Ala501Val | 5 | 28,088 |
|  |  | 11113705 | C | T | c.1529C>T | p.Thr510Met | 3 | 46,813 |
|  |  | 11113743 | G | A | c.1567G>A | p.Val523Met | 1 | 140,439 |
|  |  | 11116095 | T | G | c.1588T>G | p.Phe530Val | 10 | 14,044 |
|  |  | 11116125 | G | A | c.1618G>A | p.Ala540Thr | 2 | 70,220 |
|  |  | 11116141 | G | A | c.1634G>A | p.Gly545Glu | 1 | 140,439 |
|  |  | 11116198 | A | G | c.1691A>G | p.Asn564Ser | 2 | 70,220 |
|  |  | 11116873 | C | T | c.1720C>T | p.Arg574Cys | 2 | 70,220 |
|  |  | 11116898 | T | C | c.1745T>C | p.Leu582Pro | 1 | 140,439 |
|  |  | 11116918 | G | A | c.1765G>A | p.Asp589Asn | 1 | 140,439 |
|  |  | 11116928 | G | A | c.1775G>A | p.Gly592Glu | 1 | 140,439 |
|  |  | 11116936 | C | T | c.1783C>T | p.Arg595Trp | 6 | 23,407 |
|  |  | 11116937 | G | A | c.1784G>A | p.Arg595Gln | 2 | 70,220 |
|  |  | 11116976 | C | G | c.1823C>G | p.Pro608Arg | 1 | 140,439 |
|  |  | 11120091 | G | A | c.1846-1G>A | . | 1 | 140,439 |
|  |  | 11120106 | G | T | c.1860G>T | p.Trp620Cys | 1 | 140,439 |
|  |  | 11120110 | GAT | G | c.1867_1868del | p.Ile623HisfsTer21 | 1 | 140,439 |
|  |  | 11120143 | C | T | c.1897C>T | p.Arg633Cys | 9 | 15,604 |
|  |  | 11120144 | G | A | c.1898G>A | p.Arg633His | 1 | 140,439 |
|  |  | 11120152 | G | A | c.1906G>A | p.Gly636Ser | 3 | 46,813 |
|  |  | 11120212 | C | A | c.1966C>A | p.His656Asn | 8 | 17,555 |
|  |  | 11120370 | G | A | c.1988G>A | p.Gly663Glu | 1 | 140,439 |
|  |  | 11120408 | G | A | c.2026G>A | p.Gly676Ser | 5 | 28,088 |
|  |  | 11120436 | C | T | c.2054C>T | p.Pro685Leu | 12 | 11,703 |
|  |  | 11120441 | A | T | c.2059A>T | p.Ile687Phe | 5 | 28,088 |
|  |  | 11120442 | T | TC | c.2061dup | p.Asn688GlnfsTer29 | 1 | 140,439 |
|  |  | 11123200 | G | T | c.2167G>T | p.Glu723Ter | 1 | 140,439 |
|  |  | 11128027 | C | CA | c.2332dup | p.Arg778LysfsTer4 | 1 | 140,439 |

**Supplemental Table 3. List of variants of unknown significance (VUS) excluded from the analysis.** Genetic coordinates are mapped to GRCh38. Count refers to the number of participants having the VUS.

| **Gene** | **Chromosome number** | **Position** | **Reference allele** | **Alternate allele** | **HGVSc** | **HGVSp** | **Count** |
| --- | --- | --- | --- | --- | --- | --- | --- |
| *APOB* | 2 | 21001939 | ACTG | A | ENST00000233242:c.13480_13482delCAG | ENSP00000233242.1:p.Gln4494del | 132 |
|  |  | 21006196 | C | T | ENST00000233242:c.10672C>T | ENSP00000233242.1:p.Arg3558Cys | 299 |
|  |  | 21006239 | C | G | ENST00000233242:c.10629C>G | ENSP00000233242.1:p.Asn3543Lys | 3 |
|  |  | 21006349 | C | T | ENST00000233242:c.10519C>T | ENSP00000233242.1:p.Arg3507Trp | 1 |
|  |  | 21015387 | G | C | ENST00000233242:c.3491G>C | ENSP00000233242.1:p.Arg1164Thr | 1 |
| *PCSK9* | 1 | 55044021 | A | G | ENST00000302118:c.386A>G | ENSP00000303208.5:p.Asp129Gly | 2 |
|  |  | 55052698 | G | A | ENST00000302118:c.706G>A | ENSP00000303208.5:p.Gly236Ser | 4 |
|  |  | 55058543 | C | G | ENST00000302118:c.1399C>G | ENSP00000303208.5:p.Pro467Ala | 3 |
| *LDLR* | 19 | 11100261 | G | C | ENST00000558518.6:c.106G>C | ENSP00000454071.1:p.Asp36His | 1 |
|  |  | 11100322 | C | T | ENST00000558518.6:c.167C>T | ENSP00000454071.1:p.Ser56Phe | 1 |
|  |  | 11100328 | A | T | ENST00000558518.6:c.173A>T | ENSP00000454071.1:p.Glu58Val | 2 |
|  |  | 11100340 | C | T | ENST00000558518.6:c.185C>T | ENSP00000454071.1:p.Thr62Met | 10 |
|  |  | 11102720 | A | T | ENST00000558518.6:c.247A>T | ENSP00000454071.1:p.Ile83Phe | 1 |
|  |  | 11105262 | G | C | ENST00000558518.6:c.356G>C | ENSP00000454071.1:p.Gly119Ala | 1 |
|  |  | 11105337 | C | T | ENST00000558518.6:c.431C>T | ENSP00000454071.1:p.Pro144Leu | 1 |
|  |  | 11105379 | C | T | ENST00000558518.6:c.473C>T | ENSP00000454071.1:p.Ser158Phe | 1 |
|  |  | 11105414 | G | A | ENST00000558518.6:c.508G>A | ENSP00000454071.1:p.Asp170Asn | 22 |
|  |  | 11105415 | AC | GC | ENST00000558518.6:c.509delinsG | ENSP00000454071.1:p.Asp170Gly | 1 |
|  |  | 11106580 | G | A | ENST00000558518.6:c.710G>A | ENSP00000454071.1:p.Arg237His | 10 |
|  |  | 11106593 | C | A | ENST00000558518.6:c.723C>A | ENSP00000454071.1:p.Phe241Leu | 3 |
|  |  | 11106601 | C | G | ENST00000558518.6:c.731C>G | ENSP00000454071.1:p.Ser244Cys | 1 |
|  |  | 11106639 | C | T | ENST00000558518.6:c.769C>T | ENSP00000454071.1:p.Arg257Trp | 2 |
|  |  | 11107472 | A | G | ENST00000558518.6:c.898A>G | ENSP00000454071.1:p.Arg300Gly | 1 |
|  |  | 11111538 | A | C | ENST00000558518.6:c.1085A>C | ENSP00000454071.1:p.Asp362Ala | 60 |
|  |  | 11111558 | G | A | ENST00000558518.6:c.1105G>A | ENSP00000454071.1:p.Val369Met | 3 |
|  |  | 11111609 | G | T | ENST00000558518.6:c.1156G>T | ENSP00000454071.1:p.Asp386Tyr | 5 |
|  |  | 11113278 | G | T | ENST00000558518.6:c.1187G>T | ENSP00000454071.1:p.Gly396Val | 1 |
|  |  | 11113287 | C | T | ENST00000558518.6:c.1196C>T | ENSP00000454071.1:p.Ala399Val | 1 |
|  |  | 11113292 | CTCTTC | CTCTTG | ENST00000558518.6:c.1206delinsG | ENSP00000454071.1:p.Phe402Leu | 1 |
|  |  | 11113362 | C | T | ENST00000558518.6:c.1271C>T | ENSP00000454071.1:p.Pro424Leu | 3 |
|  |  | 11113374 | A | C | ENST00000558518.6:c.1283A>C | ENSP00000454071.1:p.Asn428Thr | 1 |
|  |  | 11113409 | A | G | ENST00000558518.6:c.1318A>G | ENSP00000454071.1:p.Arg440Gly | 4 |
|  |  | 11113561 | TCTCTTCCTA | TCTCTTACTA | ENST00000558518.6:c.1391delinsA | ENSP00000454071.1:p.Ser464Tyr | 2 |
|  |  | 11113625 | G | T | ENST00000558518.6:c.1449G>T | ENSP00000454071.1:p.Trp483Cys | 1 |
|  |  | 11113751 | T | G | ENST00000558518.6:c.1575T>G | ENSP00000454071.1:p.Asp525Glu | 6 |
|  |  | 11113762 | G | T | ENST00000558518.6:c.1586G>T | ENSP00000454071.1:p.Gly529Val | 1 |
|  |  | 11116101 | T | C | ENST00000558518.6:c.1594T>C | ENSP00000454071.1:p.Tyr532His | 1 |
|  |  | 11116132 | T | A | ENST00000558518.6:c.1625T>A | ENSP00000454071.1:p.Ile542Asn | 1 |
|  |  | 11116205 | C | G | ENST00000558518.6:c.1698C>G | ENSP00000454071.1:p.Ile566Met | 1 |
|  |  | 11116885 | G | A | ENST00000558518.6:c.1732G>A | ENSP00000454071.1:p.Val578Ile | 2 |
|  |  | 11116914 | C | G | ENST00000558518.6:c.1761C>G | ENSP00000454071.1:p.Ser587Arg | 4 |
|  |  | 11116949 | T | C | ENST00000558518.6:c.1796T>C | ENSP00000454071.1:p.Leu599Ser | 4 |
|  |  | 11116970 | C | A | ENST00000558518.6:c.1817C>A | ENSP00000454071.1:p.Ala606Asp | 14 |
|  |  | 11120454 | C | T | ENST00000558518.6:c.2072C>T | ENSP00000454071.1:p.Ser691Leu | 4 |
|  |  | 11120484 | G | T | ENST00000558518.6:c.2102G>T | ENSP00000454071.1:p.Gly701Val | 1 |
|  |  | 11120507 | A | G | ENST00000558518.6:c.2125A>G | ENSP00000454071.1:p.Arg709Gly | 1 |
|  |  | 11123315 | C | T | ENST00000558518.6:c.2282C>T | ENSP00000454071.1:p.Thr761Met | 11 |
|  |  | 11128062 | C | A | ENST00000558518.6:c.2366C>A | ENSP00000454071.1:p.Ala789Asp | 1 |
|  |  | 11129553 | G | C | ENST00000558518.6:c.2430G>C | ENSP00000454071.1:p.Trp810Cys | 1 |
|  |  | 11129573 | A | T | ENST00000558518.6:c.2450A>T | ENSP00000454071.1:p.Asn817Ile | 1 |
|  |  | 11129582 | G | A | ENST00000558518.6:c.2459G>A | ENSP00000454071.1:p.Ser820Asn | 1 |
|  |  | 11129633 | A | G | ENST00000558518.6:c.2510A>G | ENSP00000454071.1:p.His837Arg | 18 |
|  |  | 11129653 | G | A | ENST00000558518.6:c.2530G>A | ENSP00000454071.1:p.Gly844Ser | 1 |
|  |  | 11131299 | G | C | ENST00000558518.6:c.2566G>C | ENSP00000454071.1:p.Glu856Gln | 1 |

**Supplementary Table 4. UK Biobank participant characteristics post imputation of missing values stratified by familial hypercholesterolaemia (FH) carriership.** The p-values shown in the table are from the Kruskal-Wallis Rank Sum test for continuous variables, and from the Man-Whitney U test for binary variables. IQR = interquartile range; BMI = body mass index; CHD = coronary heart disease; LDL-C = low-density lipoprotein cholesterol; HDL-C = high-density lipoprotein cholesterol; CVD = cardiovascular disease; PGS = polygenic score.

|  | **Mutation negative** | **Mutation positive** | **p-value of differences** |
| --- | --- | --- | --- |
| n | 139291 | 488 |  |
| Sex (male) (%) | 63382 (45.5) | 207 (42.4) | 0.187 |
| Age (median [IQR]) | 58.0 [51.0, 63.0] | 58.0 [51.0, 63.0] | 0.803 |
| Townsend deprivation index (median [IQR]) | -2.4 [-3.8, 0.0] | -2.2 [-3.7, 0.1] | 0.367 |
| BMI, kg/m2 (median [IQR]) | 26.7 [24.1, 29.8] | 27.1 [23.9, 29.8] | 0.647 |
| Smoking status (%) |  |  | 0.827 |
| Non-smoker | 79618 (57.2) | 281 (57.6) |  |
| Former smoker | 51177 (36.7) | 173 (35.5) |  |
| Light smoker (<10 cigarettes/day) | 2021 (1.5) | 7 (1.4) |  |
| Moderate smoker (10-19 cigarettes/day) | 3497 (2.5) | 13 (2.7) |  |
| Heavy Smoker (>20 cigarettes/day) | 2978 (2.1) | 14 (2.9) |  |
| Alcohol consumption (%) |  |  | 0.492 |
| Prefer not to answer | 88 (0.1) | 1 (0.2) |  |
| 1/day | 29719 (21.3) | 93 (19.1) |  |
| 3-4 times/week | 34015 (24.4) | 135 (27.7) |  |
| 1-2 times/week | 36823 (26.4) | 130 (26.6) |  |
| 1-3 times/month | 15498 (11.1) | 54 (11.1) |  |
| Special occasions | 14383 (10.3) | 45 (9.2) |  |
| Never | 8765 (6.3) | 30 (6.1) |  |
| Family history of CHD (%) | 67013 (48.1) | 306 (62.7) | <0.001 |
| Systolic blood pressure, mmHg (median [IQR]) | 136.5 [125.0, 149.5] | 135.0 [124.5, 148.5] | 0.109 |
| Diastolic blood pressure, mmHg (median [IQR]) | 82.0 [75.0, 89.0] | 81.0 [74.0, 87.0] | 0.024 |
| Hypertension (%) | 7946 (5.7) | 35 (7.2) | 0.195 |
| Statin use (%) | 18139 (13.0) | 165 (33.8) | <0.001 |
| LDL-C PGS, mmol/L (median [IQR]) | 3.7 [3.5, 3.9] | 3.7 [3.5, 3.9] | 0.652 |
| **Blood biomarkers** |  |  |  |
| LDL-C (unadjusted for statin use), mmol/L (median [IQR]) | 3.5 [3.0, 4.1] | 3.9 [3.2, 4.8] | <0.001 |
| LDL-C (adjusted for statin use), mmol/L (median [IQR]) | 3.7 [3.1, 4.2] | 4.4 [3.7, 5.4] | <0.001 |
| HDL-C, mmol/L (median [IQR]) | 1.4 [1.2, 1.7] | 1.4 [1.2, 1.7] | 0.199 |
| Total cholesterol, mmol/L (median [IQR]) | 5.7 [4.9, 6.4] | 6.0 [5.1, 7.2] | <0.001 |
| Lipoprotein(a), nmol/L (median [IQR]) | 17.9 [9.8, 55.3] | 21.3 [12.3, 53.1] | 0.223 |
| Apolipoprotein A1, g/L (median [IQR]) | 1.5 [1.4, 1.7] | 1.5 [1.3, 1.7] | <0.001 |
| Apolipoprotein B, g/L (median [IQR]) | 1.0 [0.9, 1.2] | 1.1 [1.0, 1.4] | <0.001 |
| Triglycerides, mmol/L (median [IQR]) | 1.5 [1.1, 2.2] | 1.3 [0.9, 1.9] | <0.001 |
| C-reactive protein, mg/L (median [IQR]) | 1.3 [0.7, 2.7] | 1.2 [0.6, 2.4] | 0.045 |
| Aspartate aminotransferase, um (median [IQR]) | 24.4 [21.0, 28.8] | 25.2 [21.0, 29.5] | 0.089 |
| Alanine aminotransferase, um (median [IQR]) | 20.1 [15.4, 27.3] | 20.2 [15.6, 27.4] | 0.830 |
| Alkaline phosphatase, um (median [IQR]) | 80.1 [67.1, 95.5] | 80.6 [66.5, 95.8] | 0.571 |
| **Disease prevalence & incidence** |  |  |  |
| CHD prevalence (%) | 3890 (2.8) | 40 (8.2) | <0.001 |
| CHD incidence (%) | 5370 (3.9) | 32 (6.6) | 0.003 |
| CVD prevalence (%) | 5686 (4.1) | 45 (9.2) | <0.001 |
| CVD incidence (%) | 9038 (6.5) | 46 (9.4) | 0.011 |
| Type 2 diabetes prevalence (%) | 3593 (2.6) | 11 (2.3) | 0.757 |
| Type 2 diabetes incidence (%) | 4948 (3.6) | 19 (3.9) | 0.776 |

**Supplementary Table 5. The variables and coefficients retained by LASSO regression for monogenic FH prediction.** BMI = body mass index; CHD = coronary heart disease; LDL-C = low-density lipoprotein cholesterol; HDL-C = high-density lipoprotein cholesterol; PGS = polygenic score; Apo-A1 = apolipoprotein A1; CRP = C-reactive protein; ALT = Alanine aminotransferase.

|  | **Coefficients** |
| --- | --- |
| **(Intercept)** | -6.061379 |
| **LDL-C** | 0.252485 |
| **BMI** | -0.005819 |
| **Statin use** | 0.381582 |
| **Diastolic blood pressure** | -0.082472 |
| **Apo-A1** | -0.282415 |
| **Triglycerides** | -0.600269 |
| **CRP** | -0.004564 |
| **ALT** | -0.017213 |
| **LDL-C PGS** | -0.190587 |
| **Family history of CHD** | 0.161401 |
| **Prevalent type 2 diabetes** | -0.002958 |
| **Age x LDL-C PGS** | -0.169897 |
| **LDL-C x LDL-C** | 0.520575 |
| **Statin use x LDL-C** | 0.314738 |

**Supplementary Table 6.** **The variables and coefficients retained by LASSO regression for monogenic FH prediction excluding the LDL-C PGS.** The c-statistic of the independent test dataset was equal to 0.76 (95% CI: 0.71; 0.82).

|  | **Coefficients** |
| --- | --- |
| **(Intercept)** | -6.014496 |
| **Age** | -0.071679 |
| **Statin use** | 0.180699 |
| **Systolic blood pressure** | -0.005447 |
| **Diastolic blood pressure** | -0.060420 |
| **Apo-A1** | -0.258628 |
| **Triglycerides** | -0.561805 |
| **Family history of CHD** | 0.146255 |
| **LDL-C x LDL-C** | 0.626778 |
| **Statin use x LDL-C** | 0.406210 |

**Supplementary Figure 1. Workflow of the generation of the LDL-C PGS, FH case ascertainment and testing versus training data split of the UK Biobank’s White British participants.** The data was split according to the availability of whole-exome sequencing data. VUS = variants of uncertain significance; FH = familial hypercholesterolaemia; QC = quality control; PGS = polygenic score.

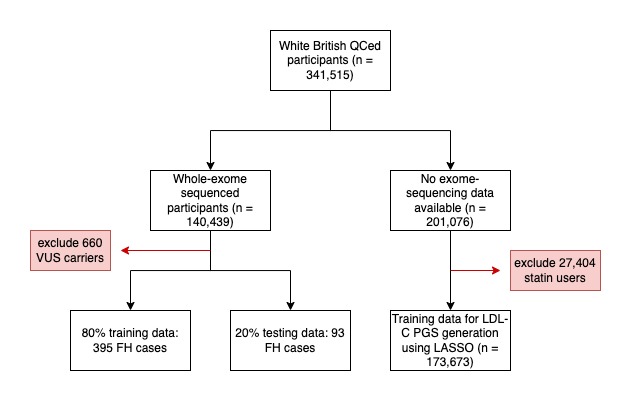

**Supplementary Figure 2. Correlation plot of the variables tested in the LASSO regression model for the prediction of monogenic FH.** The data shown here is from the training dataset as this was used to evaluated highly correlated variables prior to running the LASSO regression. LDL-C = low-density lipoprotein cholesterol; HDL-C = high-density lipoprotein cholesterol; CHD = coronary heart disease; T2D = type 2 diabetes; CRP = C-reactive protein; BMI = body mass index; ALT = alanine transaminase; SBP = systolic blood pressure; DBP = diastolic blood pressure; PGS = polygenic score; Apo-A1 = apolipoprotein A1; Lp(a) = lipoprotein A; ApoB = apolipoprotein B; AST = aspartate aminotransferase; ALP = alkaline phosphatase.

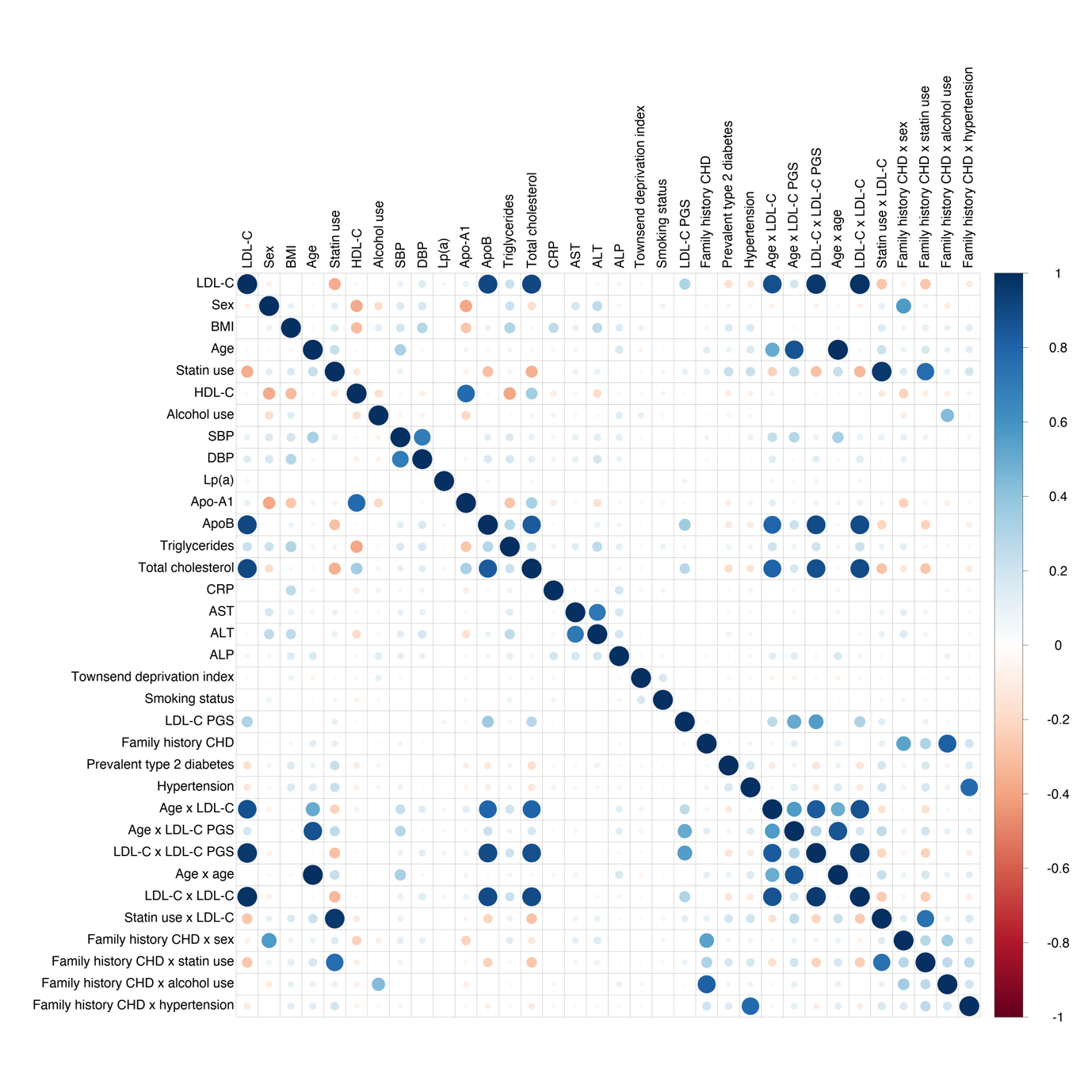

**Supplementary Figure 3. LASSO regression model feature selection and importance for monogenic FH prediction.** Feature importance is ordered by value of log odds ratio (OR) per standard deviation (SD). LDL-C = low-density lipoprotein cholesterol; CHD = coronary heart disease; T2D = type 2 diabetes; CRP = C-reactive protein; BMI = body mass index; ALT = alanine transaminase; DBP = diastolic blood pressure; PGS = polygenic score; Apo-A1 = apolipoprotein A.

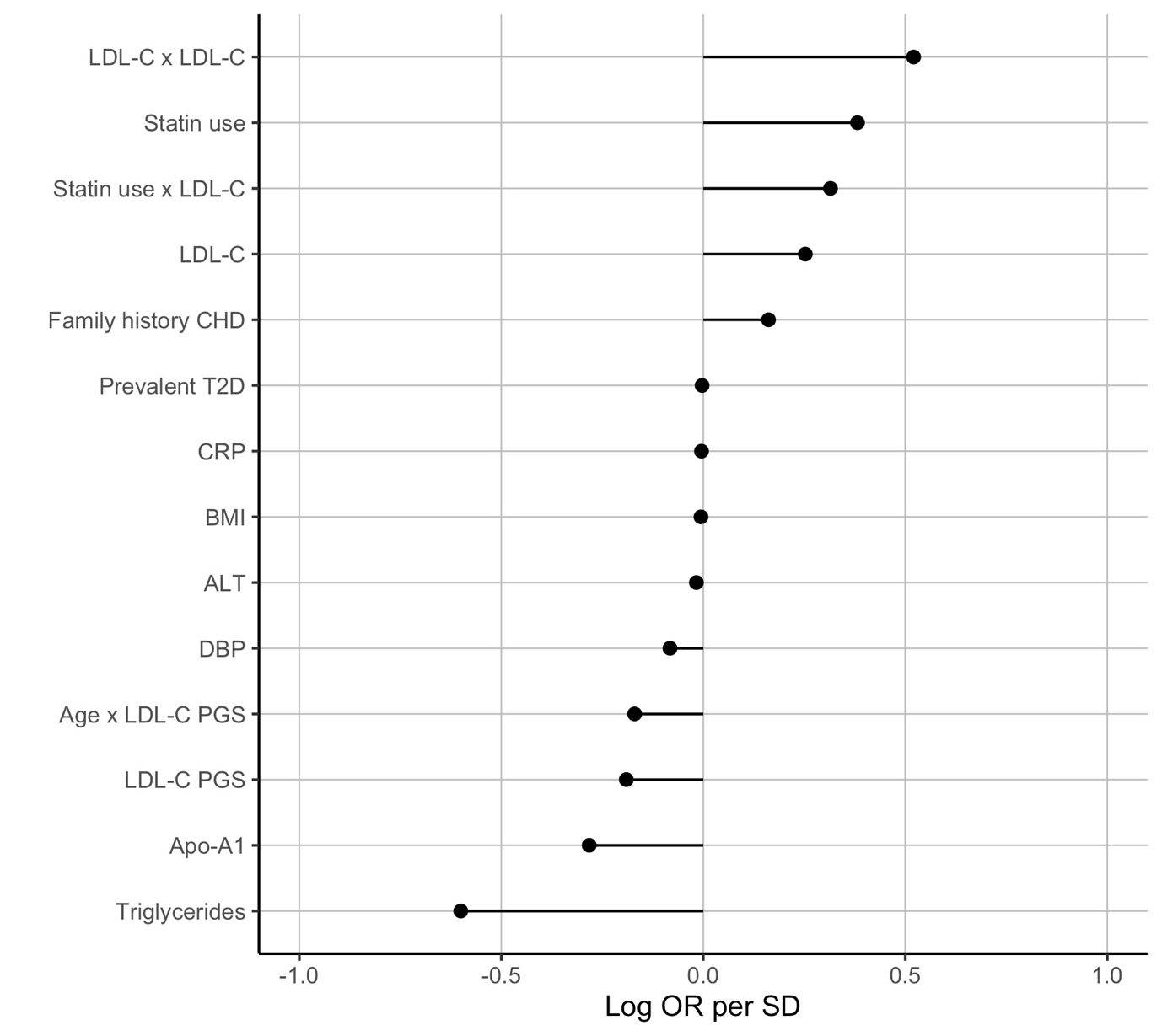

**Supplementary Figure 4. The non-linear associations of variables for the prediction of monogenic FH retained by the LASSO model.**

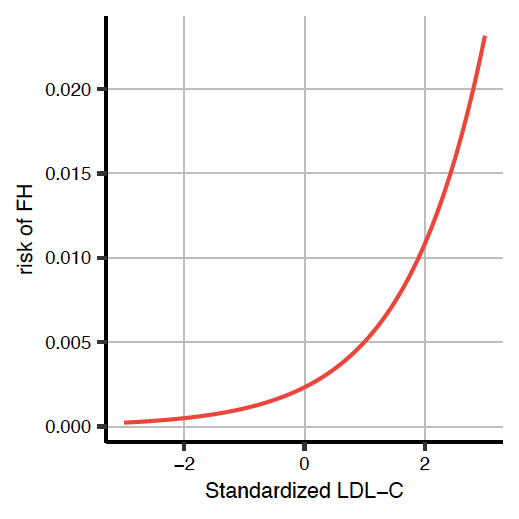

**Supplementary Figure 5. The two-stage population screening strategy for monogenic FH.** Stage 1 screen identifies individuals with a predicted probability by the LASSO model above a pre-specified threshold value, followed by a second stage of exome-sequencing. FH cases detected following this two-stage screen are brought forward for cascade testing of first-degree relatives. The sensitivity and false positive rate of the first stage depends on the threshold value chosen for the model. We assume perfect discrimination in the second stage of exome-sequencing (sensitivity of 100% and false positive rate of 0%). Cascade testing is expected to yield 1.5 additional FH cases detected for every FH case identified through the two-stage population screening strategy, as described in the results section.

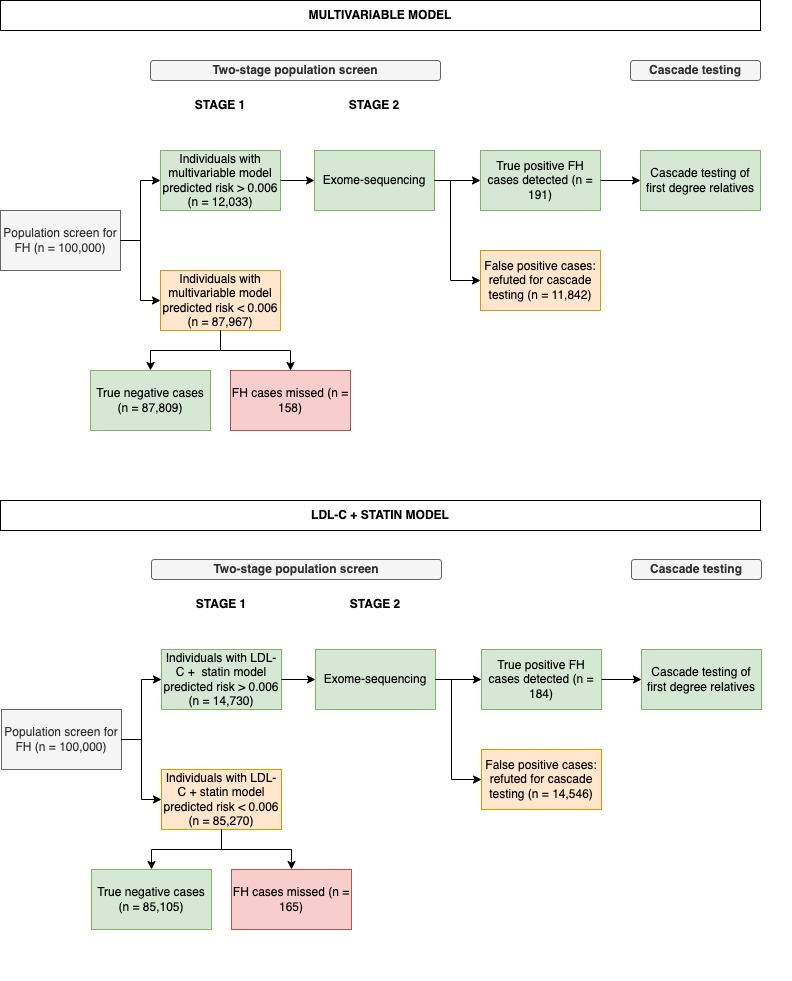
